# A Cybernetics-based Dynamic Infection Model for Analyzing SARS-COV-2 Infection Stability and Predicting Uncontrollable Risks

**DOI:** 10.1101/2020.03.13.20034082

**Authors:** Wenlei Xiao, Qiang Liu, Ji Huan, Pengpeng Sun, Liuquan Wang, Chenxin Zang, Sanying Zhu, Liansheng Gao

## Abstract

Since December 2019, COVID-19 has raged in Wuhan and subsequently all over China and the world. We propose a Cybernetics-based Dynamic Infection Model (CDIM) to the dynamic infection process with a probability distributed incubation delay and feedback principle. Reproductive trends and the stability of the SARS-COV-2 infection in a city can then be analyzed, and the uncontrollable risks can be forecasted before they really happen. The infection mechanism of a city is depicted using the philosophy of cybernetics and approaches of the control engineering. Distinguished with other epidemiological models, such as SIR, SEIR, etc., that compute the theoretical number of infected people in a closed population, CDIM considers the immigration and emigration population as system inputs, and administrative and medical resources as dynamic control variables. The epidemic regulation can be simulated in the model to support the decision-making for containing the outbreak. City case studies are demonstrated for verification and validation.

## 1. Introduction

The spread speed of SARS-CoV-2 has been emergently challenging many cities in China, especially those cities in Hubei province (Wuhan, Huanggang, Xiaogan, etc.) ^[1]^. During the currently still happening disaster, people and governments have gradually enhanced the strength of responses. Three Chinese cities (Wuhan, Huanggang and Ezhou) were shut down on Jan 23, 2020 to contain the rapidly-spreading virus; In most of Chinese cities, The first-level public health emergency response was activated on Jan 24, 2020; Huoshenshan and Leishenshan hospitals were built in less than two weeks to admit and treat patients at the epicenter of the virus; After Feb 2, 2020, Wuhan start to convert gymnasiums and exhibition centers into temporary shelter(fangcang) hospitals to accept and quarantine patients with mild symptoms. Tens of thousands of doctors and nurses from all over China have been sent to Wuhan. It seams we have always been pushed into a defensive position, that the responses were always later than the developing epidemic status. As a spontaneously summoned team, we have continuously followed the progress of this epidemic, and believe that we have found a novel forecast model to solve this problem. Base on this model, corresponding responses could be activated before the uncontrollable risks really happen.

Recently, control engineering has been maturely applied in industry. It has successfully proved its ability in analyzing the complex mechanism in a physical system. However, the control engineering has seldom been used in modeling the transmission pattern of the epidemic. Comparing its basic mechanism with an instable system, we found the dynamic infection process of COVID-19 in a city could be depicted as a cybernetic model with a positive feedback and multiple delays, and the system instability is the most issue that people and the government concern. In contrast to other classic transmission models like SIR/- SEIR (the susceptible, [exposed], infectious and recovered model), which are mostly described by a set of ordinary differential equations ^[2]^, the feedback system is constituted by a chain of discrete function blocks, such as proportional, time delayed, integral(or accumulative) blocks, and positive/negative feedback or feed-forward loop, etc. Dynamic manipulations, such as quarantining activities, medical supplies, etc., are also considered in the model. In those consideration, the model is named as Cybernetics-based Dynamic Infection Model (CDIM).

According to the daily reported numbers of confirmed and suspected COVID-19 cases from the National Health Commission of China ^[3]^, we found that cities may have non-ignorable differences on their *R*_0_. After the administrative activities carried out in most Chinese cities, such as the first-level response, the fangcang hospitals, etc., *R*_0_ also fluctuates in time significantly. Normally, those dynamic and nonlinear impacts are hard to be considered in classical transmission models (SIR, SEIR, etc.) with ordinary differential equations. In addition, the dynamic considerations can derive polymorphic city-oriented models for analyzing the uncontrollable risks in different cities with higher precision and dexterity. After several days of validation, we found the model can successfully forecast the epidemic trends in most of cities. Potential usage of this model could be a warning system for faster activating responses and predicting the shortage of medical supplies.

## 2. Review of models

In this section, we are going to discuss the disadvantages of SIR/SEIR models in epidemic forecasting, and explain how the cybernetics-based model can solve those problems.

### 2.1. Problems within SIR/SEIR

Nowadays, SIR/SEIR and their extended or modified versions are the most commonly used models to describe the spread of disease. In a closed population, the models (shown in Fig. 1) track the number of people in each of the following categories:

**Figure 1:**
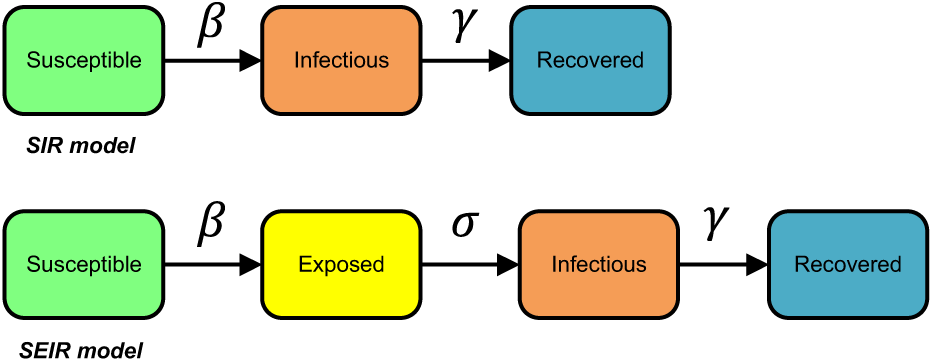
The SIR/SEIR models.

- Susceptible: Individual is able to become infected.
- Exposed: Individual has been infected with a pathogen, but due to the pathogens incubation period, is not yet infectious.
- Infectious: Individual is infected with a pathogen and is capable of transmitting the pathogen to others.
- Recovered: Individual is either no longer infectious or removed from the population.

Wherein, the infection rate *β* controls the rate of spread, the incubation rate *σ* is the rate of latent individual becoming infectious, and the recovery rate *γ* is determined by the average duration of infection. There are several disadvantages in the SIR/SEIR models:

1. Immigrated and emigrated populations are not considered in the model. Nevertheless, the traffic is far more convenient than ever before in China, so it has a significant impact on the spread.
2. The models are usually written as ordinary differential equations(ODE). It is not concise enough to help human comprehensively model the nonlinear dynamics in detail.
3. The models are relatively too simple to introduce the time-variant and probabilistic variables into the models.
4. All the parameters should be identified for estimation. In the early stage, the shortage of data makes the models infeasible to exactly forecast the trend of spread.
5. Emergency responses and medical supplies are not considered in the model, so the prediction of status has no direct recommending value to the administrative activities.
6. The parameter identification in the models strongly relies on the fidelity of real data. When the reported data have a severe deviation (like in Wuhan), it will easily lead to a forecasting failure.

In fact, what we really need to forecast is the severe situations that would happen in the future, especially at the early stage when data are in serious shortage. For example, if the short of hospital beds could be foreseen on Feb 26, 2020, the fangcang hospitals could be established much earlier. Due to the aforementioned disadvantages, SIR/SEIR cannot answer those questions.

### 2.2. Basic Principle of CDIM

In the view of modeling, there exists indeed far more advanced theories and methods in the field of control engineering. Its development is mainly benefited from the industry revolution, so that it has become a ubiquitous technology in nearly all automatic machines. In the 1940s, contemporary cybernetics began as an interdisciplinary study connecting wide fields of control systems, electrical network theory, mechanical engineering, logic modeling, evolutionary biology and neuroscience. It certainly could be employed to model the spread of epidemic. Fig. 2 gives the comparison between the current modeling methods in use for epidemic and mechanical systems. Obviously, it can be seen that the cybernetics-based modeling methods can provide more diversity in describing a system. Not only can the system be better depicted, but also can the modeling work be significantly simplified.

**Figure 2:**
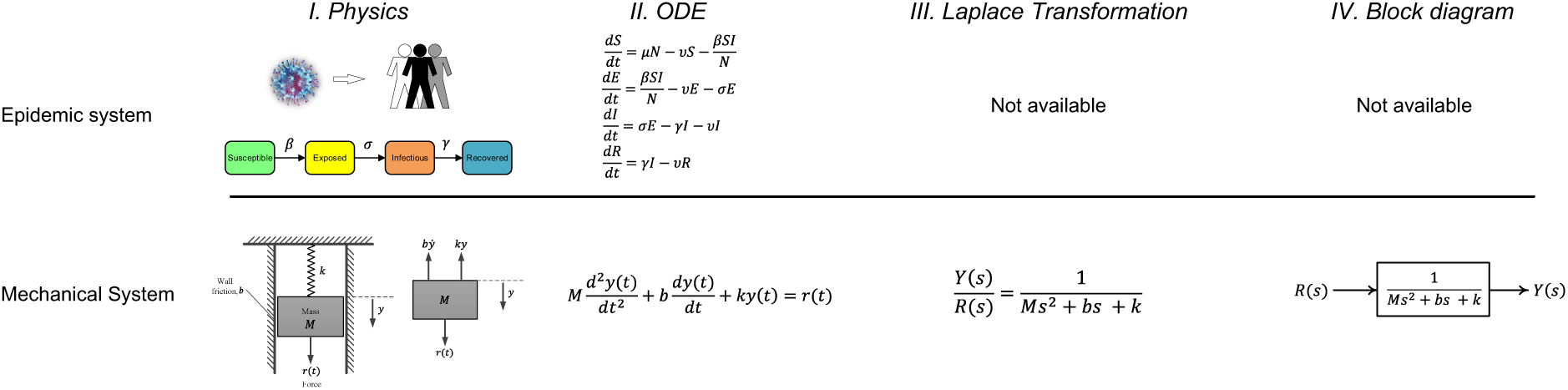
Current modeling methods in use for epidemic and mechanical systems.

In 2003, a cybernetics-based model had been firstly proposed by the authors (Ji Huan and Qiang Liu), and successfully forecasted the spread of SARS in Beijing. Fig. 3 shows the SARS model that was used to estimate the confirmed cases in Beijing and the corresponding result. The spread of SARS epidemic can be represented in a very concise manner. The positive feedback path emulates the rapid reproductive process of the epidemic, while the negative feedback represents the regulation effect from hospital (all patients are isolated at the moment of symptom onset). The result is give in Fig. 3(b), the final confirmed cases fit well with the forecasted curve. At the early stage when data were not enough, the model still could work, and the missing cases could be estimated.

**Figure 3:**
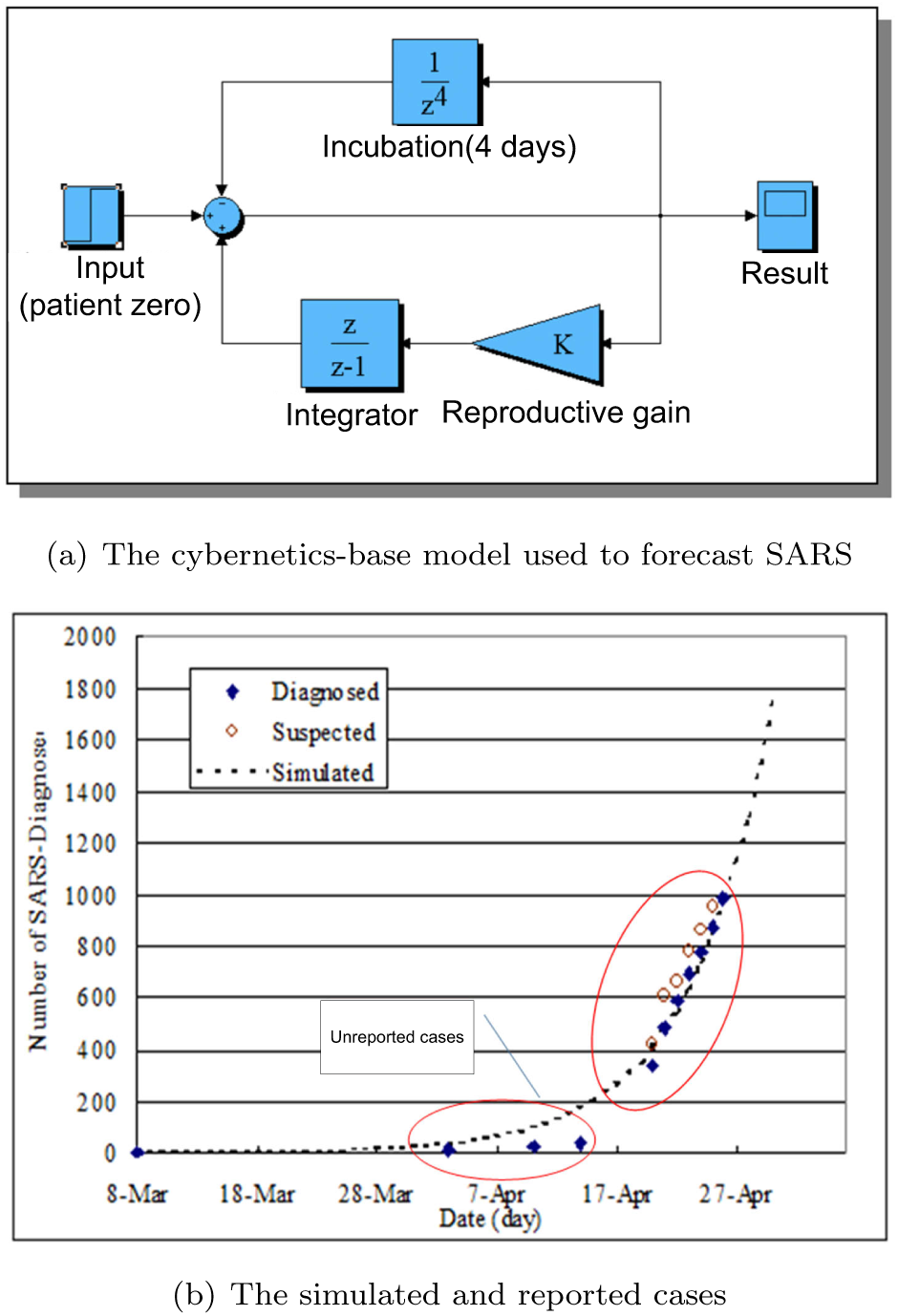
The estimation of SARS in Beijing (2003.04)

Nonetheless, there exists several basic differences in between SARS and SARS-COV-2:

1. SARS-COV-2 is contagious even in the incubation (SARS was not).
2. The traffic must be carefully depicted in the model.
3. The epidemic in multiple cities needs be forecasted.

In views of those differences, we improved the SARS model to CDIM, as shown in Fig. 4. The infection from immigration or the origin source (i.e. the Huanan Seafood Wholesale Market in Wuhan) is taken as the system input. The infection main loop generates the rapid reproduced infection. The equivalent infection rate *C*_0_ indicates the infected cases per virus carrier per day. The contact tracing rate 1 −*C*_1_ represents the effect of contact tracing of patients. The incubation bypass emulates the process of symptom onset. Since the incubation period *T*_1_ follows a Poisson distribution, the factor λ should be identified (see Eq. 1).

**Figure 4:**
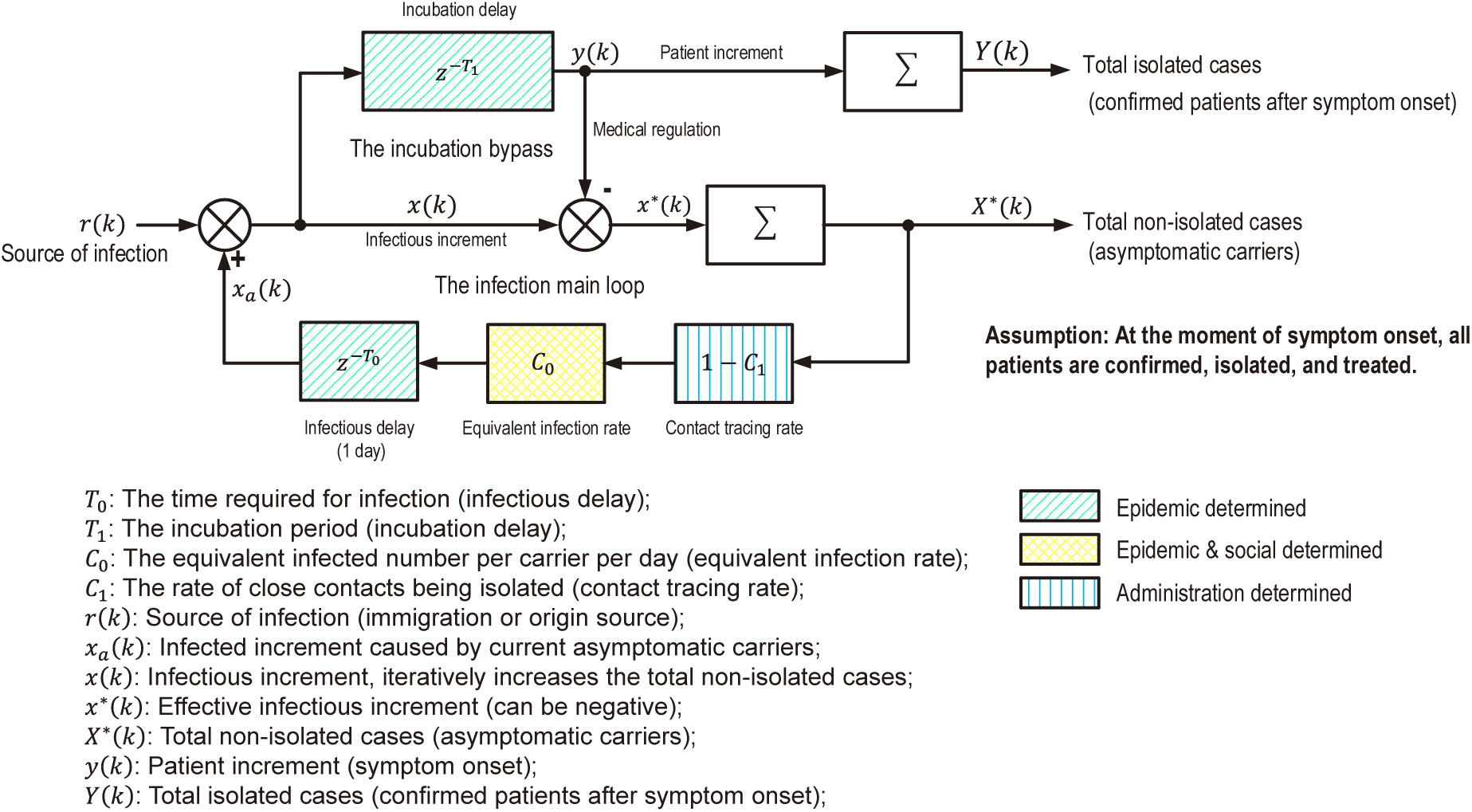
Basic principle of the cybernetics-based dynamic infection model.

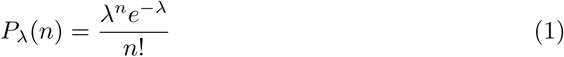

The medical regulation functions as a negative channel to control the incremental numbers of infection in the main loop. The basic model in Fig. 1 is based on a theoretical assumption, that all the patients are isolated at the moment of symptom onset (after the incubation period). When people start to wear masks and voluntarily stay indoors, *R*_0_ (namely *C*_0_) decreases accordingly. In the responsive condition, when a patient is confirmed, certain close contacts that are still in the asymptomatic incubation period would also be isolated. We introduce *C*_1_, named the contact tracing rate, to reflect this effect. When *C*_1_ ≈ 0, that means no contact tracing is imposed, the system could easily fall into an instable state if *R*_0_ ≥ 1.0. The infection model is discretized with a cyclic period of one day. In summary, *T*_0_ and *T*_1_ are mostly determined by the epidemic, *C*_0_ is determined by both epidemic and the social stress, and *C*_1_ is determined majorly by administrative activities. Eq. 2 gives the difference equations of the basic CDIM.

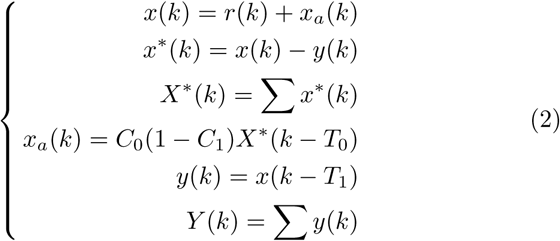

Even with the qualitative analysis, some conclusions could be made. For example, as SARS-COV-2 is contagious in its incubation, it is spreading much faster than SARS and more hard to be controlled. Consequently, experiences from SARS should not be simply copied to control the current status. This is very important issue at the early stage that should be but was not emphasized in Hubei province.

## 3. Extended city-oriented models

In China, there exists two types of city-oriented models. A typical city of the first type is Shanghai. It has enough medical supplies to admit all the patients with symptom onset, and original infectious cases were imported from immigration. Wuhan is of the second type. It was facing a serious shortage of medical supplies. Especially, the number of hospital beds in Wuhan is far less than the real needs. Moreover, due the limitation of diagnosis, there exist a confirming delay of 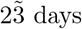. All those phenomenons should be considered in the two types of models. As the models are strongly featured by cities, we name the two types of models as Shanghai model and Wuhan model.

### 3.1. Shanghai Model

At the beginning, when the novel coronavirus from Wuhan causes concern in public, delayed and missed detection may exist in the reported number of cases. It brings troubles in estimating the epidemiological parameters and epidemic predictions ^[4,5]^. Most estimated *R*_0_ ranges from 1.5 to 4.0 ^[1,6,7,8]^. As of Jan 29, 2020, the first investigated incubation period from patients was reported, which had a mean of 5.2 days(95% confidence interval [CI], 4.1 to 7.0) and followed a Poisson distribution, and the basic reproductive number was estimated to be 2.2 (95% CI, 1.4 to 3.9) ^[6]^. However, the early sampled 425 patients had a median age of 59 years and 56% were male. Given those bias on the samples, the estimated epidemiological parameters may have deviations that might lead to great errors in the simulation. In the view of this, we used the data from Shanghai, a relatively well controlled city, to identify and calibrate the key parameters of the incubation period and the basic reproductive number. Subsequently, those parameters were used to evaluate the status of other cities (except for those cities in Hubei province). In Shanghai Model, there is no worry about the shortage of medical supplies, so a negative summation channel performs a direct control effect on the positive feedback infection loop, which is thus of paramount importance in reducing the number of total infectious cases. Two factors in the system can be regulated by the administration, *C*_0_ and *C*_1_. The confirmation delay does not affect the infecting process, but brings a direct hinder in inspecting the real number of confirmed cases. The input of the Shanghai Model mainly comes from the inspected cases from Wuhan, so the number of daily imported cases should be given by *R*_1_*x*_1_, where *R*_1_ is the infection rate of immigration, and *x*_1_ is the daily immigrated population from Wuhan. On Jan 23, 2020, Wuhan went into lockdown to contain the outbreak of the epidemic. Since then, the input from immigration was switched off. Those sudden events changed the system dynamics dramatically, and is difficult to be estimated or approximated by other transmission models. Other similar cities can reuse the Shanghai Model with some modification on the parameters *C*_0_ and *C*_1_. Nonetheless, the

Poisson distribution of incubation periods should not be changed in other cities, for the patients were derived from the same pathogeny. Comparing the reproductive speeds in different cities (as shown in Fig. 5), we found that different cities may have different reproductive gain *C*_0_(1− *C*_1_), which may depend on the generation of imported epidemic and local administrative activities.

**Figure 5:**
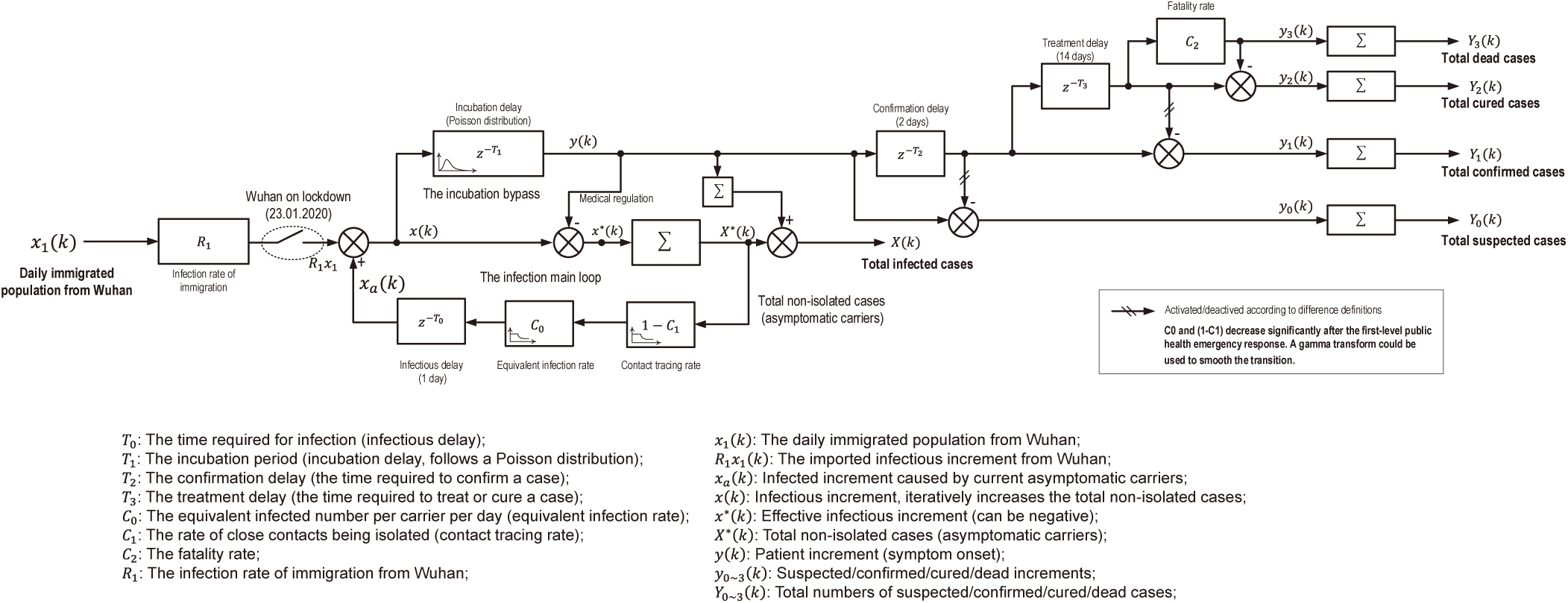
Shanghai Model.

### 3.2. Wuhan Model

There are major differences between the city Wuhan and other cities such as Shanghai, Beijing, etc. Firstly, Wuhan was facing a serious shortage of medical supplies against the outbreak of SARS-COV-2. Consequently, we designed an integral saturation module on the incubation bypass, whose capability is mostly limited by the total hospital beds; Secondly, since the original source of the coronavirus locates in Wuhan, the system input should be replaced with a spontaneous infection source; Thirdly, 5 million population had been exported outside Wuhan until Jan 23, 2020. The model should be supplemented by this emigration, as it took a large proportion of the total population (5 million left versus 9 million remained). For those sakes, Wuhan Model is designed as Fig. 6. From the daily reported news, we collected all the numbers of hospital beds with dates, including those in designated, makeshift, Huoshenshan and Leishenshan hospitals. Those numbers of beds with dates were employed in the module, admission capability of hospitals, so that the dynamic consumption of medical resources could be emulated. The system input is replaced with a spontaneous infection source, which can be simplified as a unit impulse signal, while its exact date should be deduced from later confirmed infected cases. Until the city went into lockdown, emigration had exported a number of infected patients. The factor *R*_2_ represents the infection rate at kth day of the city population *M* (*k*), which were decreasing iteratively and evenly by the daily emigration *x*_2_(*k*). The iterative formulas of *R*_2_(*k*) and *M* (*k*) are given as follow:

**Figure 6:**
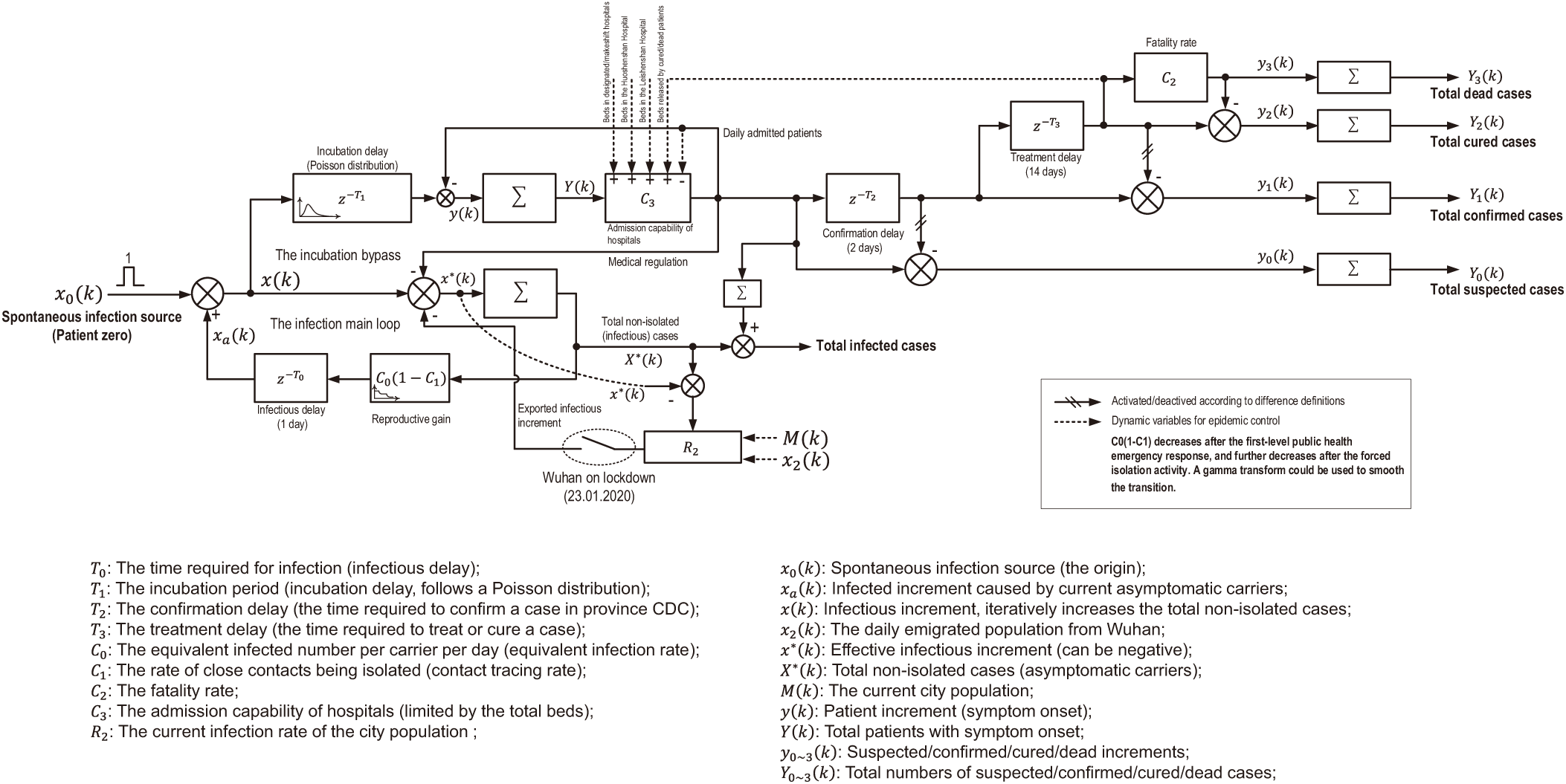
Wuhan Model.

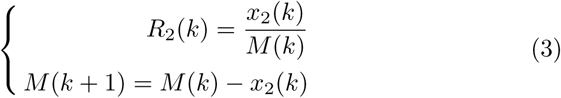

The exported infected cases are subtracted from the infection main loop.

We figured out several ways to determine the parameters in the Wuhan Model. Obviously, the incubation period can be introduced from Shanghai, while *C*_0_ and *C*_1_ should be approximated by real data. Other data such as the input and emigration were derived from collected news and reports.

## 4. Simulation case studies

Before simulation, there are basically three parameters need to be identified in the models: The Possion distributed incubation (namely λ), the reproductive gain *K* = *C*_0_(1− *C*_1_), and the traffic data. The traffic data could be derived from big data systems, while λ and *K* should be identified from real data. Note that, the reproductive gain *K* is related to the reproductive number *R*_0_:

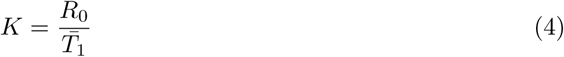

where, 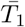 is the mean incubation.

### 4.1. Case study: Shanghai

We demonstrate the simulation result of Shanghai in Fig. 8. As of Jan 24, 2020, according to the international confirmed cases ^[9]^ and the international flights of Wuhan Tianhe Airport ^[10]^, we estimated the mean ratio of infection in the emigration from Wuhan (*R*_1_) was 0.0246%. The intercity traffic from Wuhan to Shanghai is searched from Baidu map big data. After simulation, the fitted curve of Shanghai is given in Fig. 4. The Poisson distribution factor λ of the incubation period was estimated to be 5.5 (as shown in Fig. 7), and the basic reproductive number *R*_0_ in Shanghai was 2.5 (before the first-level response) and 0.55 (after the first-level response, 95% CI, 0.48 to 0.59). Provided the latest *R*_0_ is stabilized, the final number of infected cases in Shanghai was estimated to be 344 (95% CI, 311 to 378).

**Figure 7:**
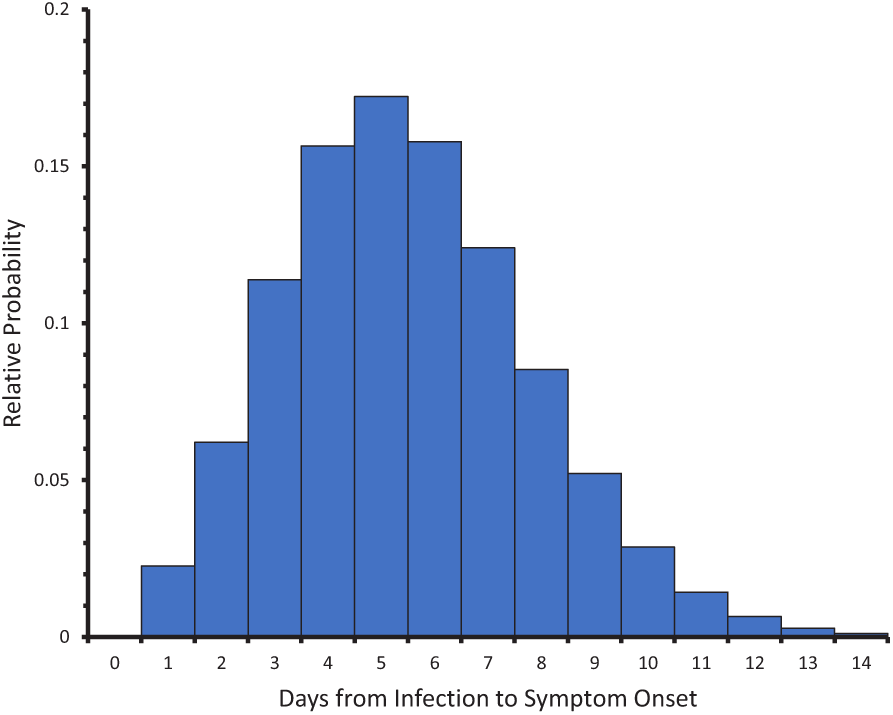
The Poisson distribution of incubation.

**Figure 8:**
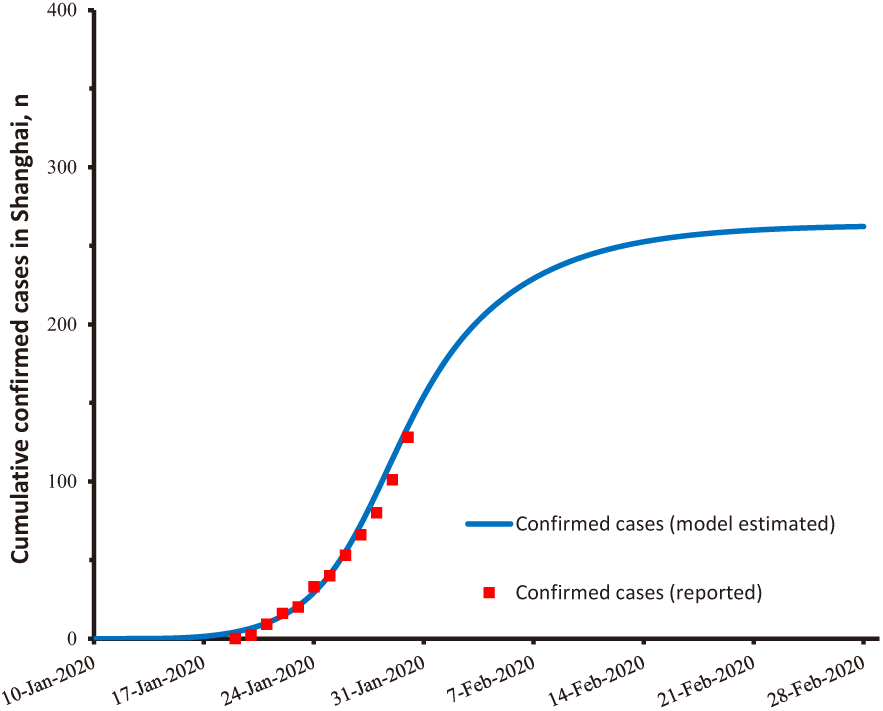
Case study: Shanghai (as of Jan 30, 2020)

### 4.2. Case study: Beijing

With the parameters identified from Shanghai, Beijing was firstly tested to evaluate its risk of stability on Feb 30, 2020. As of Feb 30 (6 days after the first-level response), it becomes clear to exactly identify the factors λ and *K*. Beijing and Shanghai are the two most important megalopolises in the main land of China, thus could share the same set of factors. In view of this, we reuse the parameters from Shanghai and deduce the fact that Beijing should postpone the end of the holiday (Chinese New Year) from Feb 2, 2020 to Feb 9, 2020. After the city resumed work, we supposed *R*_0_ would rebound to 0.8. The simulation was conducted under this assumption, and Fig. 9 reveals the difference between these two setups. The results show that the postpone of resuming work could significantly reduce the incremental infected numbers in Beijing (77.3%). This case study shows the capability of the model to support the decision-making for controlling the epidemic in a city.

**Figure 9:**
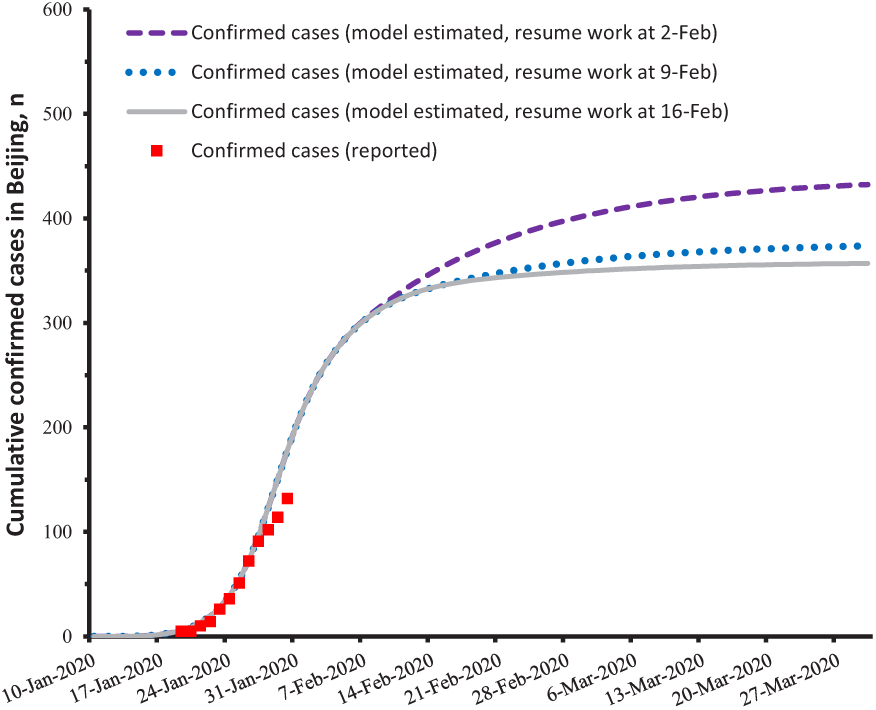
Case study: Beijing (as of Jan 31, 2020)

### 4.3. Case study: Wenzhou

Wenzhou is another featured city that was found particular with a relatively high *R*_0_, up to 4.5 (as shown in Fig. 10). Deeper investigations were taken, and it was found that the imported infectious cases from Wuhan to Wenzhou were mostly from Huanan Seafood Wholesale Market. So we may consider Wenzhou as a sub-sample of Wuhan. This phenomenon can be thus explained, and alerts should have been announced to eliminate the in-stable risk and slow down the spread speed in Wenzhou. In contrast, the inter link between Wuhan and Wenzhou helps to observe the current status of Wuhan, whose parameters could be reused in spite of the differences in their model types. After the first-level response on Jan 23, 2020, *R*_0_ decreases significantly down to 0.5 (95% CI, 0.435 to 0.556), which means both social and administrative manipulations start to work correspondingly. Provided the latest *R*_0_ is stabilized, the final number of infected cases in Wenzhou is estimated to be 527 (95% CI, 475 to 582). From the fitted curve, we could have deduced the stability of Wenzhou even early at Feb 01, 2020.

**Figure 10:**
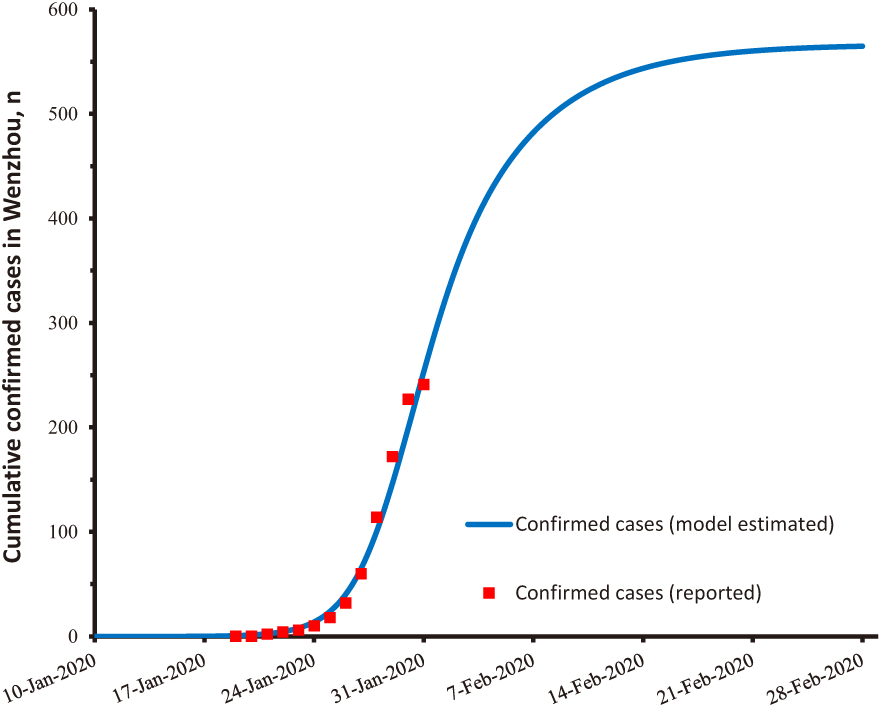
Case study: Wenzhou (as of Jan 31, 2020)

### 4.4. Case study: Wuhan

With the study of Wenzhou, it was found that the linkage between Wenzhou and Wuhan could be used to estimate the parameters in Wuhan. At that moment (2020.02.01), the status in Wuhan was extremely confusing, as a lot of infected patients were not confirmed. The hospital beds might be facing a serious shortage. Based on the collected bed data from designated, makeshift, Huoshenshan and Leishenshan hospitals, the model simulated the real situation. What can be confirmed was that emergent quarantining actions should be more strictly taken to admit the infected patients, and much more hospital beds should be prepared to contain the outbreak in Wuhan. Fig. 11 shows the simulated results. We estimated the number of infection based on the confirmed cases from the Japanese and German evacuation. There were 1.4% confirmed cases in the total evacuated population. Fig. 11(a) shows the risk of instability, and Fig. 11(b) gives the result when the forced isolation (namely the subsequent fancang hospitals) was activated since Feb 02, 2020 to curb the spread of the virus. Some days later, the forced isolation in Wuhan was implemented by those temporary shelter hospitals or fangcang. Due to the lack of data, there might exist significant errors in the simulated results. In spite of that, the epidemic stability in Wuhan was successfully estimated, and the model proves that the fancang hospitals played a vitally important role in containing the outbreak at last. The simulation of Wuhan implies a fact that, stability oriented analysis is far more important than accuracy oriented analysis. In order to make a quick decision, administrative responses could activated according to the forecasted risk of instability rather than the forecasted infected numbers. This is because the estimation of stability is usually much faster than the estimation of accuracy.

**Figure 11:**
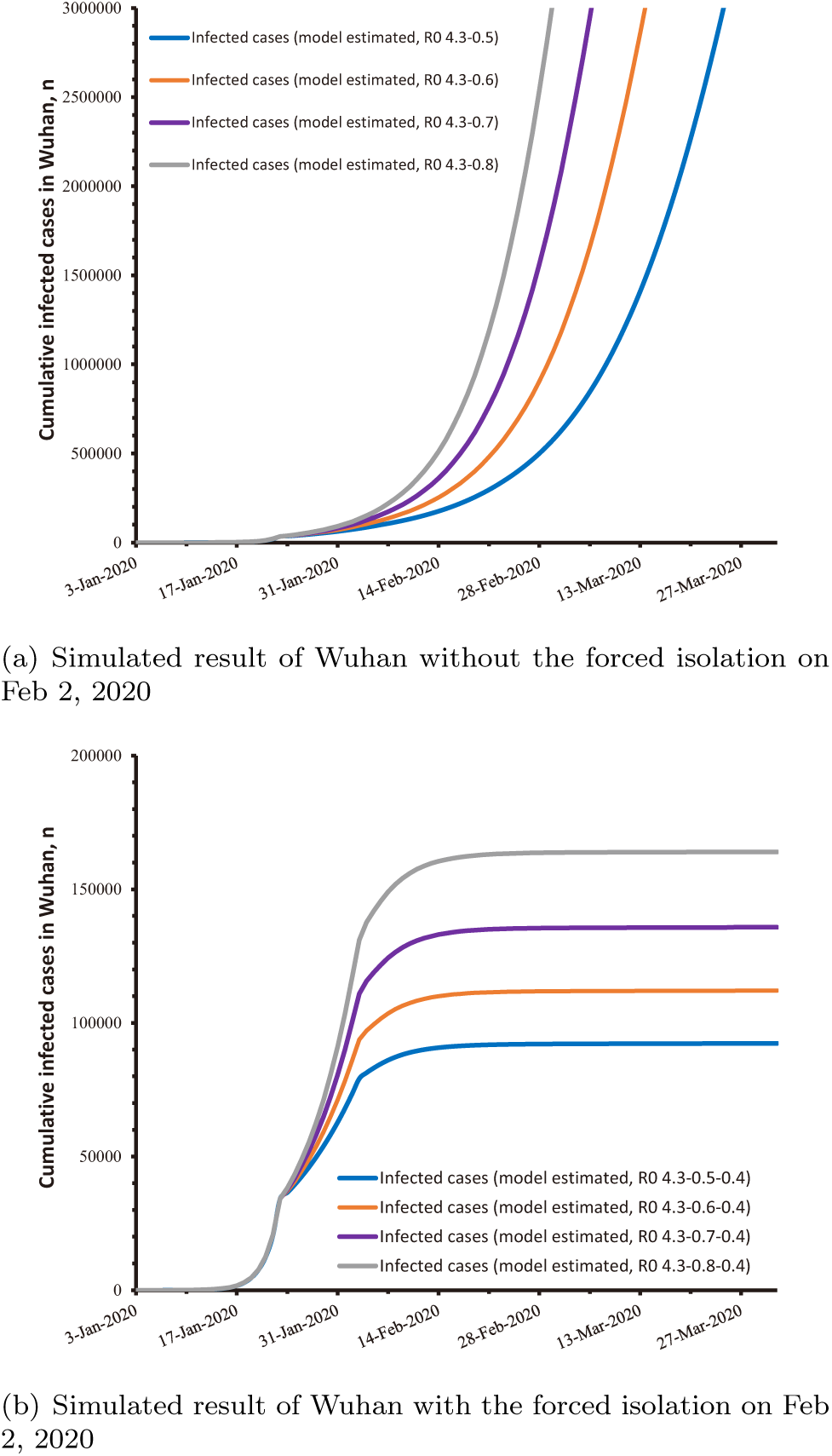
Case study: Wuhan (as of Feb 1, 2020)

**Figure 12:**
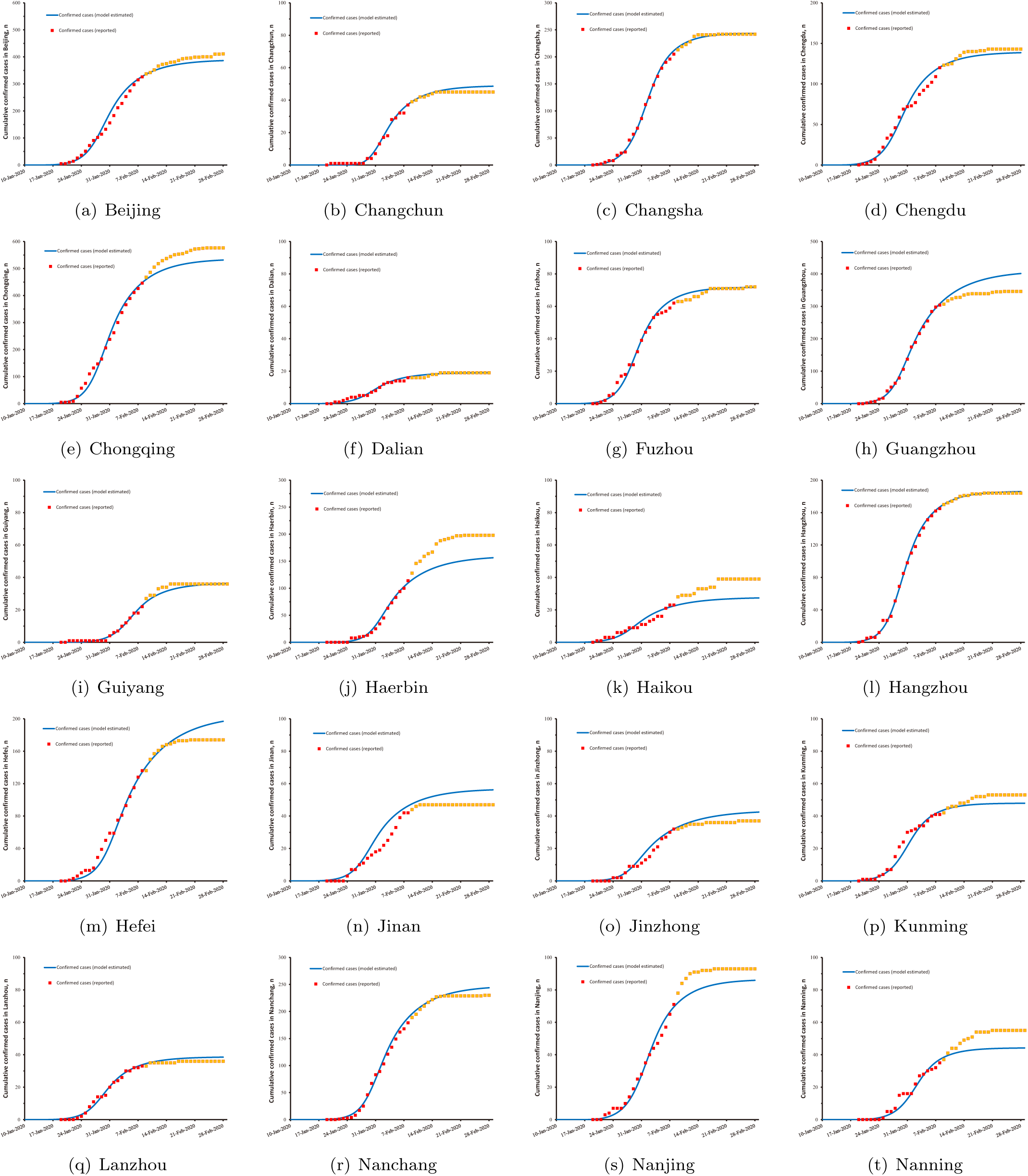
Estimation of Chinese cities, Part 1.

**Figure 13:**
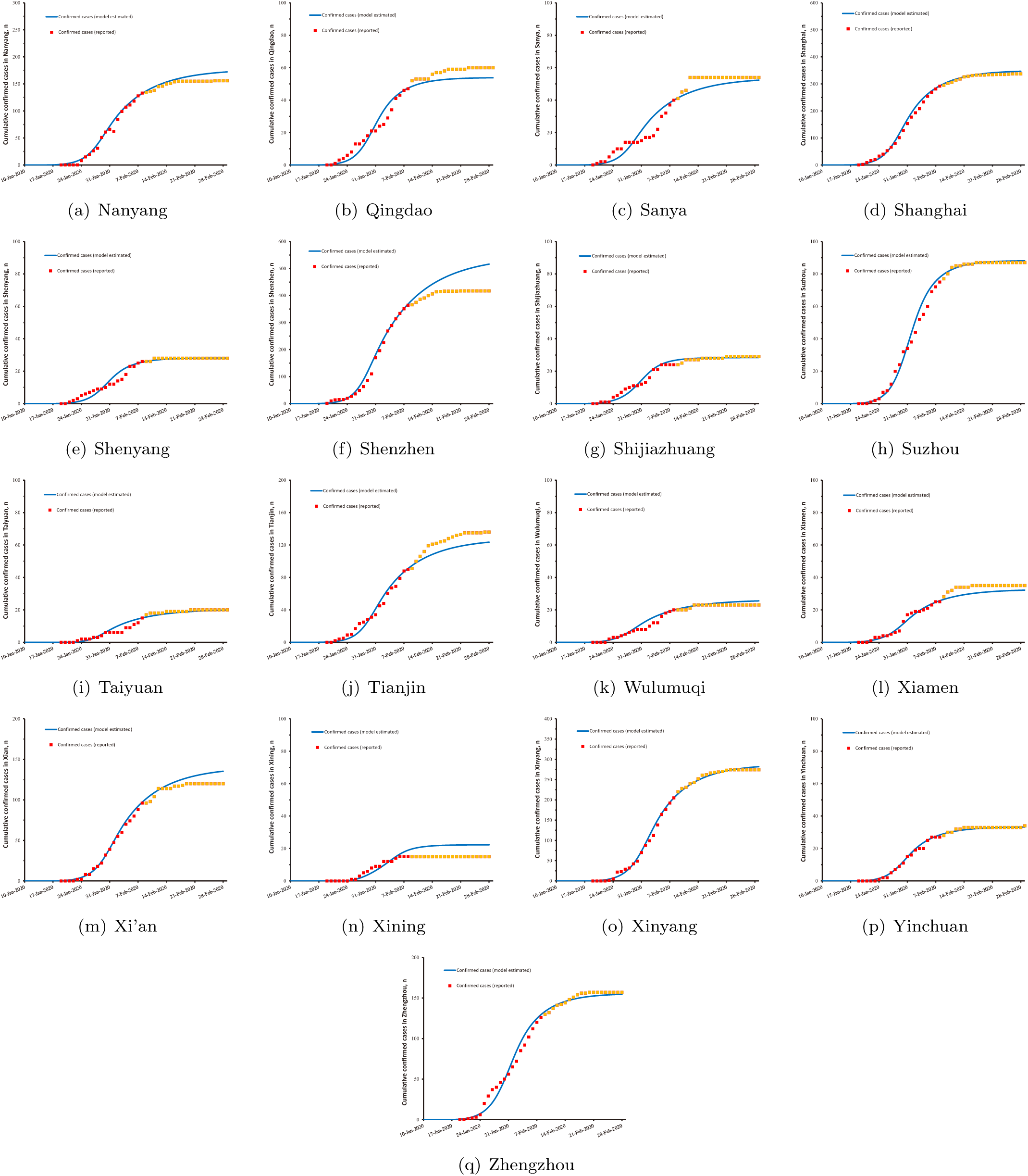
Estimation of Chinese cities, Part 2.

## 5. Conclusion and Discussion

We present a Cybernetics-based Dynamic Infection Model to simulate the epidemiologic characteristics of SARS-COV-2. Two polymorphic models for different city types are derived with considerations on their medical, immigration/emigration population and administrative conditions. Shanghai, Beijing, Wenzhou and Wuhan were studied to validate the infection models. More simulated results based on the successive data from 37 cities in China are listed in the appendix. Results demonstrate that the model can successfully simulate the dynamic infecting process of COVID-19, and uncontrollable risks in a city can be predicted before they come true. Additionally, administrative management can be verified by the model so that decisions can be made in time with more confidence according to the forecasts and warnings. Combined with big data of the intercity traffics, the research work is even more helpful to future epidemic predictions. Uncontrollable risks in multiple cities could be monitored in parallel, emergency responses could be activated faster, and medical supplies would be produced precisely and transported to the right place at the right time.

Currently, the outbreak in China is almost brought under control, while the international spread is developing rapidly. The global transmission is just like the situation in China one month ago. In view of this, many parameters could be reused to forecast the spread in foreign countries (South Korea, Italy, Iran, etc.), experiences should be learned from China to prevent more cities from instability. From the proposed model, we could infer the following conclusions:

1. Medical responsibility: Medical supplies should be prepared abundantly to admit all the patient with symptom onset;
2. Social responsibility: People should reduce contacts as much as possible, so that the factor *C*_0_ can be decreased;
3. Administrative responsibility: The contact tracing rate *C*_1_ plays a vitally significant role in stabilizing the spread.

All the above mentioned responsibilities should be emphasized, so that the current situation could be stabilized before the end of May.

## Data Availability

All raw data are collected from web.

## Acknowledgement

The group is led by Prof. Dr. Qiang LIU and Associate Prof. Dr. Wenlei XIAO. Qiang and Wenlei contribute to design the dynamic infection model based on the original cybernetic idea of modeling SARS infection, proposed by Prof. Dr. Ji HUAN in 2003. Ph.D candidates, Pengpeng SUN, Liuquan WANG and Chenxin ZANG, contribute to develop the software program, as well as to identify the model parameters and conduct the simulation. Ph.D candidates, Chenxin ZANG and Sanying ZHU, contribute to the data collection and pre-processing. Associate Prof. Dr. Liansheng GAO contributes to the confidential analysis of the predictive results. Special acknowledgement to Associate Prof. Dr. Xiangmei Chen and Prof. Dr. Wei ZHANG for their viewpoints of medicine and public health.

Also we thank the following people for their data collection and visualization work in this research: Associate Prof. Dr. Chong PENG, Dawei GAO, Jie ZOU, Jimeng LIU, Jian WANG, Changri XIONG, Chenghao ZHANG, Shiping WANG, Kang CHENG, Zihui SUN, Zhenshuo YIN, Shaoyu WU, Maocheng XU, Jinyue LI, Mingzhen XIA, Jian WANG, Junqi XU, Shuo SU, Shuang YAO, Qingshuo CHANG, Shicheng XU, etc.

## Appendix: Simulation of 37 Chinese cities (as of Feb 9, 2020)

In order to validate the CDIM model, 37 cities were simulated on Feb 9, 2020, and more data (yellow points) were used to verify the precision of estimation. Most of cities fit well with the estimation, though some cities have some deviations. This is mainly caused by the dynamic quarantining level that changes *R*_0_. It implies that, in order to better estimate the epidemic, a dynamic parameter identification algorithm should be developed in future work.

